# Development of a Real-Time Risk Tracking Model for Neurological Deterioration in Ischemic Stroke Patients Based on Blood Pressure Dynamics

**DOI:** 10.1101/2024.04.09.24305591

**Authors:** Jihoon Kang, Maeng Seok Noh, Juneyoung Lee, Youngjo Lee, Hee-Joon Bae

## Abstract

**Objectives:** Using the significant link between blood pressure fluctuations and neurological deterioration (ND) in patients with ischemic stroke, this study aims to develop a predictive model capable of real-time tracking of ND risk, enabling timely detection of high-risk periods.

**Methods:** A total of 3,906 consecutive ischemic stroke patients were recruited. As an initial predictive model, a polytomous logistic regression model, incorporating clinical parameters to estimate a probability of ND occurring within and beyond 12 hours post-stroke onset, was developed. To refine ND risk assessments over time, we subsequently introduced an iterative risk-tracking model that utilizes continuously updated blood pressure measurements. We endeavored to integrate these models, assessing their combined discriminative capacity and clinical utility, with a particular emphasis on pinpointing time periods of increased ND risk.

**Results:** ND rates were observed at 6.1% within the first 12 hours and 7.3% during the following 60 hours. We noted variations in incidence rates and their distribution over time across predefined patient groups. Significant predictors of ND varied among these subgroups and across different time periods. The iterative risk-tracking model maintained a consistent relationship between blood pressure variables and ND risk across different patient groups, successfully forecasting ND within a 12-hour window. The integrated models achieved an area under the receiver operating characteristic curve (AUC) ranging from 0.68 to 0.76. This performance effectively narrowed down the critical window for ND risk identification without sacrificing predictive accuracy across diverse patient groups. At 90% and 70% sensitivity settings, the combined model precisely identified the periods of highest ND risk, showing slightly higher or comparable specificity and positive predictive values relative to other models.

**Conclusion:** This study presents a novel approach for real-time monitoring of ND risk in ischemic stroke patients, utilizing BP trends to identify critical periods for potential intervention.

## Introduction

Ischemic strokes pose a substantial risk of secondary neurological worsening, with approximately a 30% likelihood of neurologic deterioration (ND) within the initial week following onset.^1–3^ This deterioration is attributed to a variety of heterogeneous conditions, such as hemorrhagic transformation, progression of ischemic lesions, and early recurrences,^4,5^ all of which critically affect stroke outcomes and increase mortality rates.^6–8^

ND is characterized by two notable aspects that warrant particular attention. Firstly, the risk of ND fluctuates with time elapsed since the stroke event.^9,10^ Specifically, in the first 12 hours post-stroke, the incidence of ND is 12 events per 1,000 person-hours, which then declines to 4 events in the next 12 hours and further to 2 events in the subsequent 24 hours.^9^ Secondly, there exists variability in ND risk across different clinical profiles, such as variations in stroke severity and the application of recanalization therapies.^11–14^

Given these dynamics, the use of conventional risk prediction models for monitoring and detecting ND risk faces limitations.^15,16^ Such models, which apply a static set of risk predictors to estimate overall risk at a single time point, fall short in guiding the optimal timing for interventions aimed at preventing ND.^17^ Moreover, the variability in ND risk across different clinical scenarios poses challenges to the utility of models that fail to account for this diversity.^18,19^ Although there have been efforts to create models tailored to specific scenarios, such as for patients with minor stroke and large-vessel occlusion eligible for intravenous thrombolysis,^20^ their scope of application remains limited.

In response, we have developed a novel model designed to dynamically track changes in ND risk over time, accommodating the variability of risk across different clinical settings. This model performs iterative risk estimations at specific time points and identifies high-risk intervals, utilizing patient profiles and continuous blood pressure (BP) data as indicators. BP, a critical physiological marker for both the brain and overall bodily functions,^21,22^ has a well-documented strong association with ND risk.^23–25^ This study evaluates the model’s performance and explores its practical applicability through estimating the positive predictive values in real-world settings.

## Methods

### Study Subjects and Data Collection

This research utilized clinical data from ischemic stroke patients who were admitted to Seoul National University Bundang Hospital within 48 hours of symptom onset between April 2008 and March 2015 and were registered in the multicenter prospective stroke registry in South Korea, known as the Clinical Research Collaboration for Stroke in Korea (CRCS-K).^26^ We collected comprehensive clinical information, including demographics (age, sex), medical history (hypertension, diabetes mellitus, hyperlipidemia, atrial fibrillation), and detailed stroke characteristics. These characteristics encompassed the National Institute of Health Stroke Scale (NIHSS) score, stroke mechanism, the location of occluded cerebral arteries pertinent to acute ischemic stroke, types of reperfusion therapies administered, and the achievement of successful recanalization. Data were sourced both from the registry’s database and by reviewing electronic health records (EHR).

Blood pressure (BP) data for each patient were systematically retrieved from the EHR repository and formatted to include BP values alongside their respective measurement times. BP monitoring during hospital stays was guided by the discretion of treating physicians, in alignment with stroke practice guidelines and institutional protocols. Initially, in stroke units or intensive care units, BP was monitored hourly, with adjustments tailored to individual patient needs. This schedule was later modified to 4-to 8-hour intervals upon the patient’s transfer to the general ward. BP measurements were conducted on the non-hemiparetic arm in a supine position, ensuring the patient was in a resting state, using non-invasive semi-automated devices (IntelliVue MP20; Philips Medizin System, Böblingen, Germany) and an NIBP device (BP-8800, Fukuda Denshi Co., Ltd., Japan), supplemented by traditional sphygmomanometers when necessary.

### Standard Protocol Approval and Patient Consent

The Institutional Review Board (IRB) approved the collection of clinical information for the purpose of monitoring and enhancing the quality and outcomes of stroke care. Given the anonymized nature of the data and the minimal risk posed to participants, the IRB granted a waiver for informed consent. Furthermore, the IRB authorized using the registry database for this study, including the retrospective collection of blood pressure (BP) and other necessary clinical data.

### Study Outcome

The primary study outcome was the occurrence of ND within 72 hours of hospital admission. ND was defined by meeting at least one of the following criteria: the emergence of any new neurological symptoms or signs, an increase of two or more points in the total National Institute of Health Stroke Scale (NIHSS) score, or an increase of one or more points in the NIHSS sub-scores for the level of consciousness or motor function.^9,27^

### Statistical Analysis

Our dynamic risk prediction model was developed to estimate the ND risk within 12 hours of each BP measurement, beginning from hospital admission (**Figure 1 and Supplemental Table 1**). The initial model (***Model A***) utilized polytomous logistic regression to compute ND risk within the first 12 hours of admission (***P1***) and up to 72 hours (***P2***), incorporating clinical parameters. A subsequent model (***Model B***) calculated the 12-hour ND risk iteratively, factoring in BP levels at each measurement and changes in BP from the initial and most recent measurements. This process involved adjusting the coefficients of predictors for four predetermined patient groups: those with successful recanalization, symptomatic steno-occlusion of major cerebral arteries (SSO) and small vessel occlusion (SVO), and others (**Supplemental Figure 1**).^11–14^

**Figure 1.**
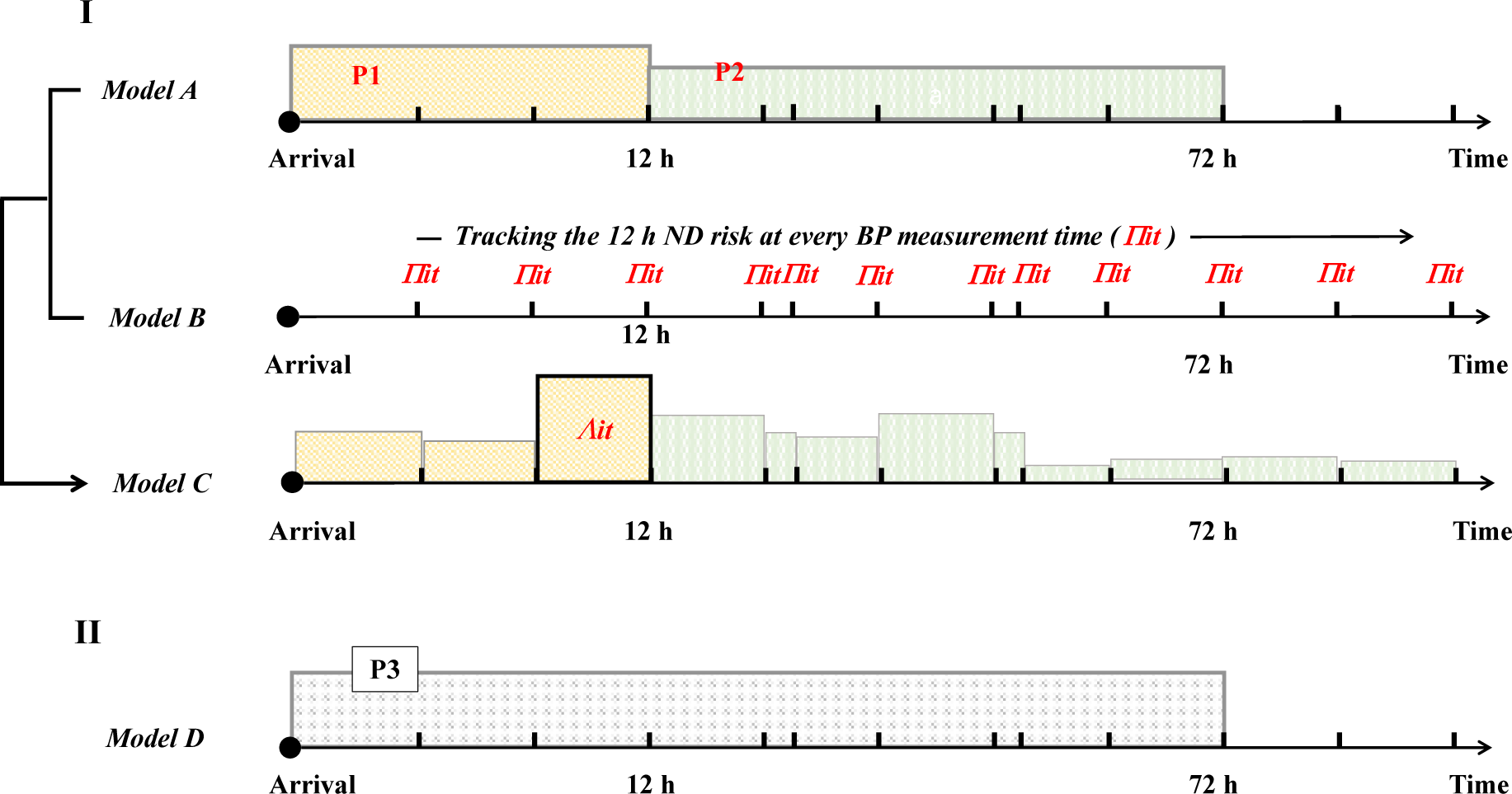
The Schematic flow of tracking the high-risk time zone. The upper panel presents the concept of risk-tracking prediction models from the hospital arrival (panel I). With clinical parameters, it predicted the ND probability within 12 h (P1) and between 12 and 72 h (P2) using the polytomous regression model (***Model A***). The second model renewed the ND probability (Π_*it*_) iteratively with BP parameters collected at every measurement time point (***Model B***). Both models were combined to estimate the change of ND probability in ***Model C*** and delineated the maximum risk time (*Λit*). The lower panel (II) shows the conventional prediction model to estimate the ND risk at once, irrespective of the risky time zone (***Model D***).

A combined model (***Model C***) was then established to identify the highest risk period using ND risk estimations from ***Models A and B***. The efficacy of ***Model C*** was assessed by measuring the area under the curve (AUC). For comparative analysis, AUC values for ***Model A*** and a conventional prediction model (***Model D***) — a binary logistic regression model for ND probability within 72 hours with clinical parameters — were calculated. Additionally, the clinical applicability of ***Model C*** was evaluated by comparing its positive predictive values (PPVs) and specificity at 90% and 70% sensitivity levels with those from ***Models A and D***. Statistical analyses were conducted using SAS version 9.4 (SAS Institute, Inc., Cary, NC).

### Data Availability Statement

Data in this study are available from the corresponding author by request.

## Results

The study population consisted of 3,904 patients with ischemic stroke, presenting an average age of 67.1 ± 13.4 years and a median National Institutes of Health Stroke Scale (NIHSS) score of 4 (interquartile range, 1–8). In alignment with the study protocol, these patients were stratified into four distinct groups, revealing significant imbalances in age, time from symptom onset to hospital arrival, baseline NIHSS score, presence of atrial fibrillation, and stroke history (**Table 1**).

**Table 1.** Baseline Characteristics of Total Study Subjects and Their Comparisons among the Predetermined 4 Patient Groups.

| Variables | Total subjects<br>(n=3904) | Stroke groups |  |  |  | *P* |
| --- | --- | --- | --- | --- | --- | --- |
|  |  | Successful<br>Recanalization<br>(n=432) | SSO<br>(n = 525) | SVO<br>(n = 659) | Other<br>(n = 2256) |  |
| Male | 2342 (60.0%) | 202 (58.2%) | 76 (52.1%) | 392 (59.7%) | 1503 (61.2%) | 0.13 |
| Age, y, mean ± SD | 67.1 ± 13.4 | 69.7 ± 11.9 | 69.0 ± 13.7 | 64.4 ± 12.7 | 66.9 ± 13.7 | <0.001 |
| Onset to arrival, h, median (IQR) | 5.4 (1.6 – 16.1) | 1.3 (0.6 – 3.0) | 4.5 (1.4 – 14.0) | 9.3 (3.2 – 22.7) | 6.1 (1.8 – 17.6) | <0.001 |
| Baseline NIHSS, median (IQR) | 4 (1 – 8) | 14 (9 – 20) | 8 (3 – 17) | 3 (1 – 4) | 3 (1 – 6) | <0.001 |
| Hypertension | 2678 (68.6%) | 306 (70.8%) | 361 (68.8%) | 456 (66.0%) | 1555 (68.9%) | 0.35 |
| Diabetes Mellitus | 1165 (29.8%) | 118 (27.3%) | 159 (30.3%) | 195 (28.2%) | 693 (30.7%) | 0.38 |
| Dyslipidemia | 1117 (28.6%) | 110 (25.5%) | 144 (27.4%) | 203 (29.4%) | 660 (29.3%) | 0.37 |
| History of stroke | 846 (21.7%) | 85 (19.7%) | 137 (26.1%) | 122 (17.7%) | 502 (22.3%) | 0.003 |
| Atrial fibrillation | 844 (21.6%) | 236 (54.6%) | 176 (33.5%) | 2 (0.3%) | 430 (19.1%) | <0.001 |
| Stroke subtypes |  |  |  |  |  | NA |
| Small vessel occlusion | 691 (17.7%) | NA | NA | 691 (100.0%) | NA |  |
| Large artery atherosclerosis | 1287 (32.9%) | 88 (20.4%) | 226 (43.0%) | NA | 973 (43.1%) |  |
| Cardioembolism | 1040 (26.6%) | 263 (60.9%) | 207 (39.4%) | NA | 568 (25.2%) |  |
| Other determined | 180 (3.6%) | 14 (3.2%) | 29 (5.5%) | NA | 137 (6.1%) |  |
| Undetermined | 708 (18.1%) | 67 (15.5%) | 63 (12.0%) | NA | 578 (25.6%) |  |
| Recanalization therapy |  |  |  |  |  | NA |
| IVT-only | 269 (6.9%) | 56 (13.0%) | 37 (7.0%) | 27 (3.9%) | 149 (6.6%) |  |
| EVT-only | 249 (6.4%) | 171 (39.6%) | 46 (8.8%) | NA | 32 (1.4%) |  |
| Combined treatment | 261 (6.7%) | 205 (47.5%) | 29 (5.5%) | NA | 27 (1.2%) |  |
| ND |  |  |  |  |  |  |
| All time window | 590 (15.1%) | 109 (25.2%) | 124 (23.6%) | 68 (9.8%) | 289 (12.8%) | < 0.001 |
| ND, arrival – 12h | 206 (5.3%) | 65 (15.0%) | 38 (7.2%) | 19 (2.7%) | 84 (3.7%) | < 0.001 |
| ND, 12 – 72h | 301 (7.7%) | 40 (9.3%) | 67 (12.8%) | 40(5.8%) | 154 (6.8%) |  |
| ND, 72h – | 83 (2.1%) | 4 (0.9%) | 19 (3.6%) | 9 (1.3%) | 51 (2.3%) |  |

Values were the number of patients (percentage) except for specific descriptions. P values were estimated by Pearson chi-square test, ANOVA test, and Kruskal Wallis test, as appropriate. Stroke groups were those with successful recanalization therapy, symptomatic steno-occlusion of major cerebral arteries (SSO), small vessel occlusion (SVO), and others. NIHSS was abbreviated for National Institute of Health for Stroke Scale, SD for standard deviation, IQR for interquartile range, IVT for intravenous thrombolysis, EVT for endovascular treatment, NA for Not applicable, ND for neurologic deterioration, ICA for internal carotid artery, and MCA for middle cerebral artery.

The analysis highlighted substantial differences in ND occurrence within 72 hours post-admission among the different patient groups. The group achieving successful recanalization exhibited the highest ND incidence at 24.5%, followed by patients with SSO at 20.0%. The others and SVO group demonstrated lower ND rates at 10.5% and 8.5%, respectively (P < 0.001). Furthermore, the study identified varying patterns of ND occurrence over time among these groups, with a notable peak in ND rates primarily within the first 12 hours, followed by a decreasing trend thereafter. This temporal distribution of ND risk is further illustrated in **Figure 2**, emphasizing the early high-risk period for ND and subsequent decline.

**Figure 2.**
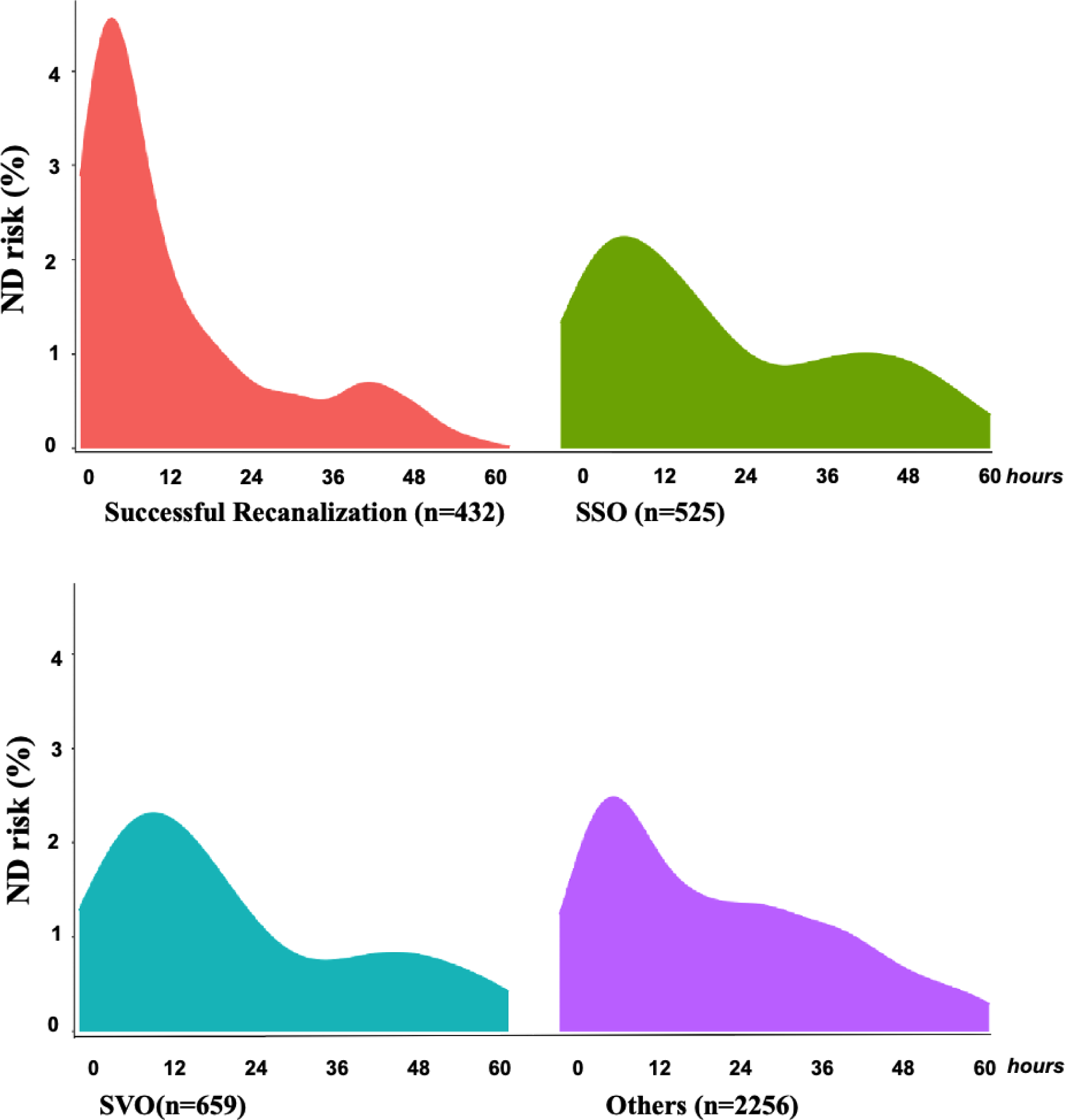
The density plots for ND over time after hospital arrival by the groups. The densities represented the risk of neurologic deterioration (ND) by predetermined groups, while the X-axis denoted the hour (h) after hospital arrival. SSO is abbreviated for symptomatic steno-occlusion of major cerebral arteries (SSO), and SVO is denoted for small vessel occlusion.

### Development of prediction models

***Model A*** was developed to estimate ND risk within the first 12 hours following admission and extending through the subsequent 60 hours through predetermined patient groups. They revealed the significant variations of predictors by group and time zone (**Table 2**). Early after onset, factors such as time from onset to hospital arrival and stroke mechanism were significantly associated with ND risk in some patient groups, whereas symptomatic vascular diseases became more relevant predictors in later hours.

**Table 2.**
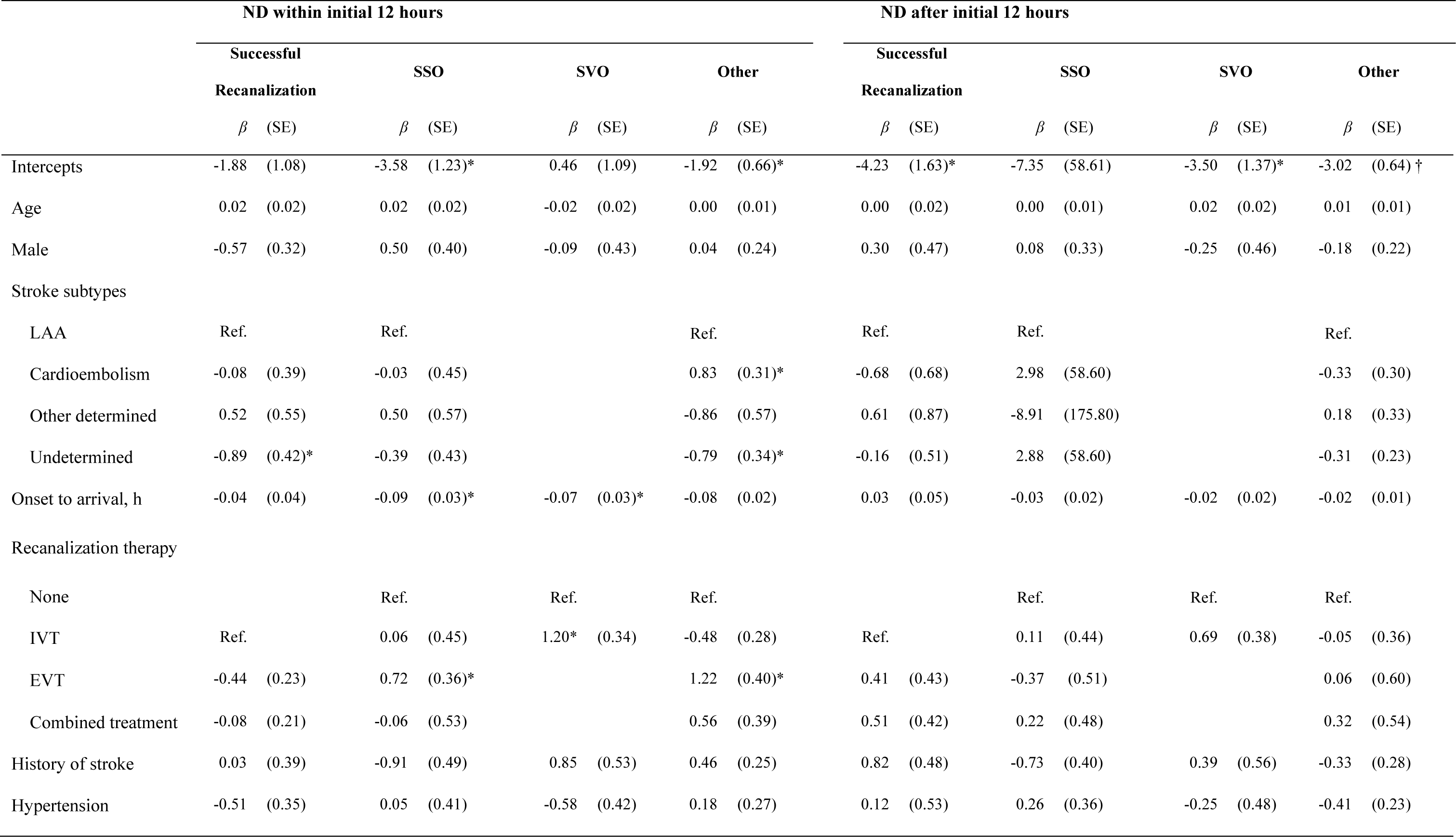
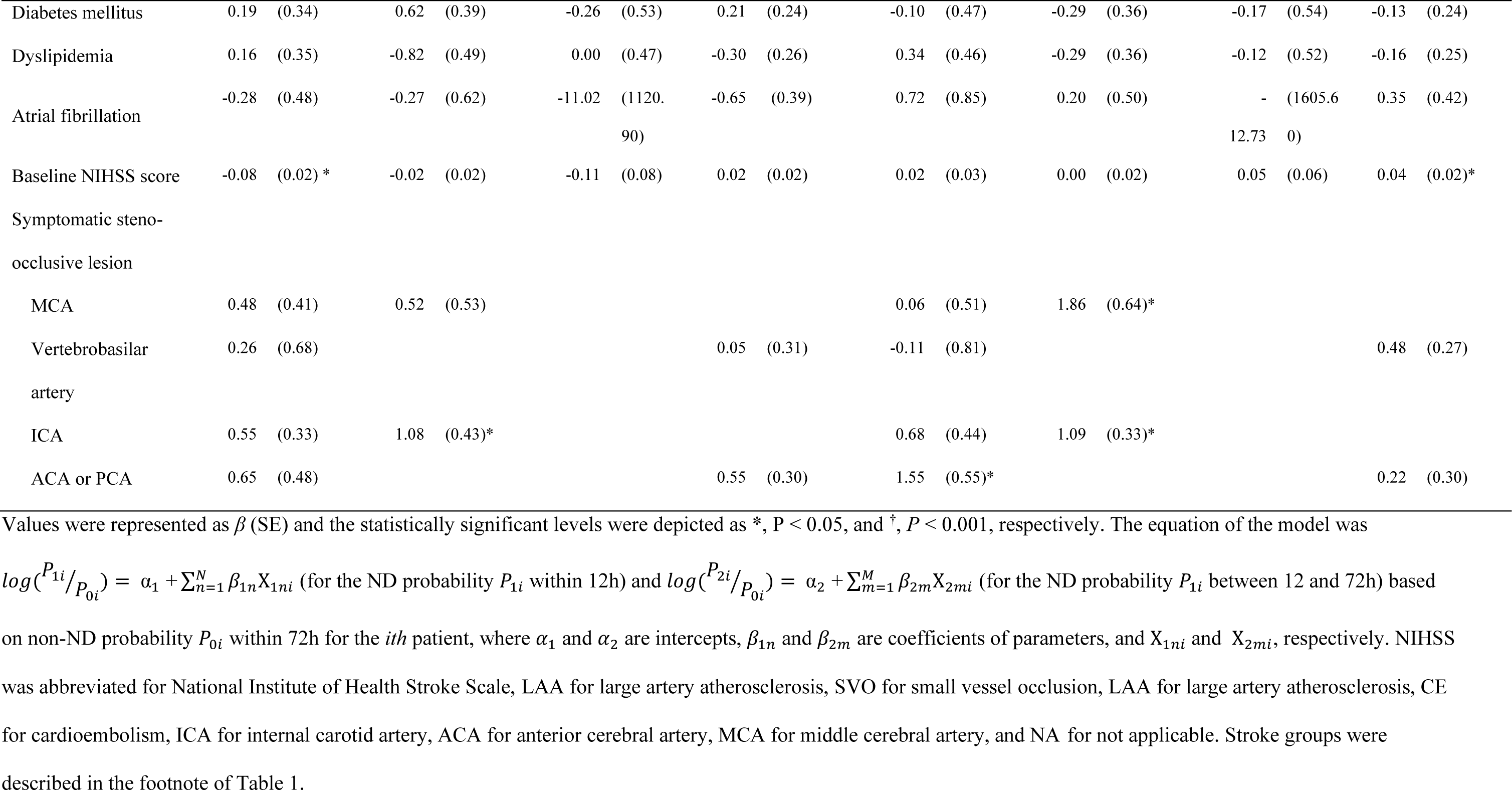
Predictors of Neurological Deterioration in the Polytomous Logistic Regression (*Model A*)

Following this, ***Model B,*** an iterative risk-tracking model at every BP measurement point, was developed using logistic regression models segmented by the four patient groups (**Table 3**). This model demonstrated consistency in the relationship between BP parameters and ND probabilities, specifically for forecasting ND risk within a 12-hour timeframe from each measurement point. ND risk within 12 hours from each measurement was associated positively with systolic BP at each time point and negatively with a BP change from baseline to each time point among all the patient groups. However, it was noted that the magnitude of these effects— effect sizes—exhibited variation across different patient groups.

**Table 3.**
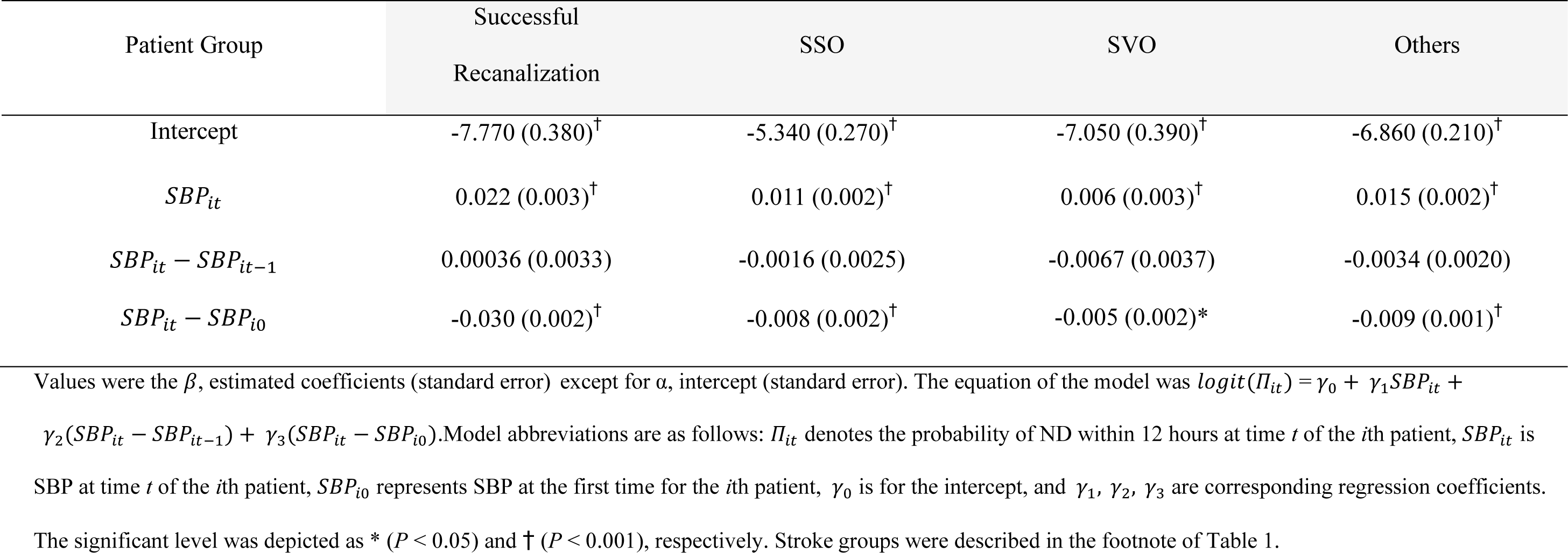
Iterative Risk-Tracking Models Using Blood Pressure Data in 4 Patient Groups (*Model B*)

| Patient Group | Successful<br>Recanalization | SSO | SVO | Others |
| --- | --- | --- | --- | --- |
| Intercept | -7.770 (0.380)<sup>†</sup> | -5.340 (0.270)<sup>†</sup> | -7.050 (0.390)<sup>†</sup> | -6.860 (0.210)<sup>†</sup> |
| $SBP_{it}$ | 0.022 (0.003)<sup>†</sup> | 0.011 (0.002)<sup>†</sup> | 0.006 (0.003)<sup>†</sup> | 0.015 (0.002)<sup>†</sup> |
| $SBP_{it} - SBP_{it-1}$ | 0.00036 (0.0033) | -0.0016 (0.0025) | -0.0067 (0.0037) | -0.0034 (0.0020) |
| $SBP_{it} - SBP_{i0}$ | -0.030 (0.002)<sup>†</sup> | -0.008 (0.002)<sup>†</sup> | -0.005 (0.002)* | -0.009 (0.001)<sup>†</sup> |

Values were the $\beta$ , estimated coefficients (standard error) except for $\alpha$ , intercept (standard error). The equation of the model was $logit(\Pi_{it}) = \gamma_0 + \gamma_1 SBP_{it} +$
$\gamma_2(SBP_{it} - SBP_{it-1}) + \gamma_3(SBP_{it} - SBP_{i0})$ . Model abbreviations are as follows: $\Pi_{it}$ denotes the probability of ND within 12 hours at time $t$ of the $i$ th patient, $SBP_{it}$ is SBP at time $t$ of the $i$ th patient, $SBP_{i0}$ represents SBP at the first time for the $i$ th patient, $\gamma_0$ is for the intercept, and $\gamma_1, \gamma_2, \gamma_3$ are corresponding regression coefficients.

***Model C***, which identified the highest-risk time zones for ND, was set up with predicted values from ***Models A and B***. On reaching the maximal predicted risk over the next 12 hours, they optimized the coefficients of predicted values, which presented wide ranges of variations across different patient groups (**Table 4**). The model’s crimination ability to distinguish between cases with and without ND in this timeframe was quantified, as shown in **Table 5**. The AUC for ***Model C*** varied between 0.68 and 0.76, demonstrating comparability to ***Models A and D*** across different patient groups. Notably, this similarity in performance was observed despite ***Model C***’s unique focus on predicting distinct outcomes, such as the high-risk time window, from simplified time zone comparison (the first 12 h vs. the subsequent 60 h, ***Model A***), and ND occurrence (***Model B***).

**Table 4.** Combined models for identifying the high-risk time zone (*Model C*)

| Patient Group | Successful<br>Recanalization | SSO | SVO | Others |
| --- | --- | --- | --- | --- |
| Intercept ($\alpha$) | -3.57 (0.39)<sup>†</sup> | -3.21 (0.54)<sup>†</sup> | -4.37 (0.53)<sup>†</sup> | -4.40 (0.27)<sup>†</sup> |
| $P_{1i} + P_{2i}$ | 5.02 (1.32)<sup>†</sup> | 5.11(1.10)<sup>†</sup> | 8.73 (1.61)<sup>†</sup> | 7.09 (1.00)<sup>†</sup> |
| $max_t (\Pi_{it})$ | 14.1 (5.90)* | 9.54 (18.90) | 35.3 (33.50) | 70.7 (18.20)<sup>†</sup> |

**Table 5.** Comparisons of Model Performance, Specificity, and Positive predictive values at 90% sensitivity level among different models.

| Patient Group<br>(No. of BP measurements) | Successful<br>Recanalization<br>(n = 41,335) | SSO<br>(n = 59,421) | SVO<br>(n = 28,626) | Others<br>(n = 142,006) |
| --- | --- | --- | --- | --- |
| **AUC** |  |  |  |  |
| *Model C* | 0.68 | 0.71 | 0.76 | 0.71 |
| *Model A* | 0.66 | 0.71 | 0.76 | 0.70 |
| *Model D* | 0.63 | 0.72 | 0.76 | 0.70 |
| **PPV** |  |  |  |  |
| *Model C* | 13% | 16% | 6.0% | 7.4% |
| *Model A* | 13% | 16% | 5.5% | 6.6% |
| *Model D* | 12% | 15% | 5.4% | 6.7% |
| **Specificity** |  |  |  |  |
| *Model C* | 28% | 31% | 37% | 37% |
| *Model A* | 25% | 30% | 32% | 28% |
| *Model D* | 17% | 28% | 30% | 29% |

The specificity and PPV were calculated with 90% sensitivity. The **Model D** was set up with the logistic regression model, while **Model A** was the polytomous regression model, and **Model C** was the composite models of predicting the maximum ND risk within using **Model A** and iterative risk tracing model (**Model B**). AUROC was abbreviated for area under the receiver operating curve and PPV for positive predictive value.
Model A
Model B
Model C
Model D

At a sensitivity level of 90%, ***Model C*** exhibited slightly higher or comparable specificity values relative to the other models, suggesting its effectiveness as a risk-monitoring tool in accurately narrowing down the high-risk time window for ND. Additionally, the PPV of ***Model C***, indicating its capacity to alert patients within the highest risk window, was marginally superior to that of the other models, further supporting its clinical relevance.

Additional analysis estimating specificity and PPV at a 70% sensitivity level revealed consistent trends, underscoring the model’s reliability and consistency in performance (**Supplemental Table 2**).

## Discussion

This study introduced an innovative iterative risk-tracing model designed to predict the high-risk time zone for ND following ischemic stroke. Our model exhibits comparable or marginally improved predictive accuracy over traditional models and, crucially, could narrow down the highest risk time zone for ND occurrence.

Recent literature increasingly underscores the importance of pinpointing the precise timing for heightened ND risk, reflecting the dynamic nature of ND in the early post-stroke period and across varied clinical contexts.^16–18,20^ While some existing models restrict their focus to the initial 24 hours post-stroke, others limit their application to specific treatments like intravenous thrombolysis or endovascular therapy.^18,28^ Our study addresses these limitations by extending the tracking time zone to account for the variable timing of ND occurrences and evolving risk profiles.

Despite similar AUC values (ranging from 0.68 to 0.76) when compared to conventional models (***Model A and D***), ***Model C,*** a combination stands out by narrowing the predictive window to 12 hours, enhancing its clinical utility by enabling preemptive alerts for ND risk—a capability not paralleled by other models.

In the context of evolving stroke treatment paradigms aiming at improving outcomes, the increasing demand on resources and the workload for continuous monitoring, particularly for patients with mild neurological symptoms, necessitates efficient monitoring tools.^7,29^ Our ND-monitoring tool addresses this need by enabling targeted monitoring for high-risk patients, potentially alleviating the burden on healthcare systems.^30^

The occurrence of ND peaks within the first 12 hours following stroke onset, diminishing subsequently. This study highlights the dynamic nature of ND risk predictors, which vary with the elapsed time from stroke onset. Consequently, accurately assessing and mitigating ND risk necessitates consideration of this temporal aspect.

Moreover, our analysis revealed distinct temporal patterns of ND risk across different patient groups. Notably, patients with small vessel occlusion exhibited a marginally higher risk of ND within the first 24 hours, with a gradual decline in risk thereafter. Conversely, patients who underwent successful recanalization demanded heightened vigilance in the initial hours post-stroke.

This variability underscores the importance of tailoring ND risk assessment and intervention strategies to individual patient profiles and the timing post-stroke, fostering a more nuanced approach to stroke management and prevention strategies.

BP parameters stand as crucial physiological markers of bodily function, readily measurable and modifiable in the stroke population.^30^ The biological rationale for, and the benefits of, frequent BP measurements are well-established.^31^ This study has corroborated the significance of BP levels and their fluctuations from initial and subsequent measurements, reinforcing the value of BP as a predictor of neurologic outcomes.

Echoing findings from previous research that underscored the predictive utility of continuously monitored vital signs for cardiac arrest,^32^ our results suggest that incorporating additional parameters, such as glucose levels, into the monitoring regimen could further enhance the accuracy of ND risk prediction in stroke patients.^33^ This approach emphasizes the potential for a more comprehensive and dynamic assessment strategy, leveraging multiple vital parameters to refine risk evaluations and intervention timing for stroke care.

Several limitations of this study warrant attention. Firstly, the development and validation of our prediction model were conducted through retrospective analysis. To confirm its validity, a prospective study is needed, which could provide more robust evidence of the model’s predictive accuracy and reliability in real-time clinical settings. Secondly, while our study utilized an integrative model to identify high-risk time zones for ND, further research is required to explore and validate various alarming criteria. This step is crucial to establish the model’s practical feasibility and optimize its application in clinical practice. Lastly, the BP data employed in our analysis were collected retrospectively. Although this approach offers benefits in terms of practical applicability, the retrospective nature of data collection limits the ability to ensure consistent BP measurement intervals and the regular assessment of ND risk. Future studies should aim to address these limitations to enhance the predictive model’s accuracy and utility in stroke care.

In conclusion, our study presents a dynamic risk-tracing model that significantly advances the predictive accuracy and clinical applicability of ND risk assessment in ischemic stroke patients. By focusing on the critical early hours post-stroke and utilizing real-time BP data, our combined model offers a novel approach to identifying high-risk patients, potentially facilitating timely interventions and improving stroke outcomes. Future studies should aim to validate these findings through prospective research, explore additional predictive parameters, and assess the model’s integration into clinical workflows. Our work lays the foundation for more personalized and efficient stroke care, emphasizing the need for continuous innovation in monitoring and intervention strategies.

## Data Availability

Data in this study are available from the corresponding author by request.

